# Enhanced Predictive Accuracy of the Revised Risk Analysis Index Over the 5-Factor Modified Frailty Index for Postoperative Outcomes in Olecranon Fractures

**DOI:** 10.1101/2025.04.06.25325335

**Authors:** Cameron J. Sabet, Dilibe Ekowa, Noah Baca, Nithin Gupta, Taylor J. Manes, Michael Kessler

## Abstract

**Objective:** To compare the predictive accuracy of the Risk Analysis Index (RAI) versus the 5-factor Modified Frailty Index (mFI-5) for postoperative outcomes in olecranon fracture open reduction internal fixation (ORIF).

**Methods:** This retrospective study analyzed 3,987 patients from the ACS-NSQIP database who underwent olecranon ORIF between 2015-2020. Outcomes included 30-day mortality, non-home discharge (NHD), complications, readmission, and extended length of stay. Predictive accuracy was assessed using area under ROC curves (AUROC).

**Results:** RAI demonstrated superior predictive accuracy for NHD (AUROC: 0.81 vs 0.68, p<0.001), major complications (AUROC: 0.72 vs 0.65, p=0.05), and reoperation (AUROC: 0.63 vs 0.57, p=0.03) compared to mFI-5. Severely frail patients identified by RAI showed significantly increased odds for NHD (OR: 4.78, p=0.005), extended length of stay (OR: 2.83, p=0.008), and major complications (OR: 9.23, p=0.03). No significant differences were found between indices for mortality, minor complications, or readmission rates.

**Conclusion:** The RAI demonstrates superior discriminatory accuracy compared to mFI-5 for predicting adverse outcomes after olecranon ORIF, particularly for NHD and major complications. Implementation of RAI in preoperative assessment may improve risk stratification and resource allocation for olecranon fracture patients.

## Introduction

Olecranon fractures can severely impact the utility, flexibility, and long-term use of the elbow. As one of the most common upper limb fractures (10% of cases), surgical intervention can lead to complications in about 33.3% of patients.^1^ With increasing aging populations generally and the incidence of osteoporosis rising, these fractures are becoming a major issue in the management of the upper limb in orthopedic practice.^2^

Surgical repair of olecranon fractures is typically considered a low-risk procedure, even for those requiring open reduction and internal fixation (ORIF). However, outcomes are determined largely by patient factors including age, frailty, and comorbidities such as lifestyle and PDH.^3^ Given an aging population and potential complications/decreased quality of life following ole was cranon ORIF, stratification tools may be employed to identify patients at risk of adverse postoperative outcomes. Frailty is defined as decreased physiologic reserve, reducing a patient’s ability to recover from stress such as surgery. Geriatric care has long been interested in the management of frailty’s physiologic and pathologic consequences. This concept has been investigated at length using newly developed tools such as frailty indexes in order to generate ever-more accurate assessments of the frailty status of the aging population within patient pools.^1–5–7^

Many surgical outcomes can now be simulated using one or more of the various frailty assessment tools developed in recent years. The Modified Frailty Index (mFI-5) has been a popular choice in the literature due to its simplicity, predictive accuracy, and ease of application across various specialties.^86–102^ However, a newer frailty index, the Risk Analysis Index (RAI) is an increasingly popular index that more directly analyzes the true state of frailty rather than its five distinct adjacent variables, four of which are a measure of comorbidity. The RAI is more comprehensive, and captures a wider spectrum of possible indicators of frailty than the five variables driving mFI-5.^11–213^ Although the mFI-5 has been utilized previously in the orthopedic literature, there is a growing body of evidence in the neurosurgical literature that supports greater predictive ability and accuracy for the RAI.^24–26^ Therefore, this study seeks to understand the utility of frailty, as measured by the mFI5 and RAI, for predicting perioperative outcomes among patients undergoing ORIF for olecranon fractures

## Methods

### Data Source

This study utilized a dataset from the American College of Surgeons National Surgical Quality Improvement Program (NSQIP), which includes a large de-identified dataset of patients undergoing surgical procedures in multiple institutions. Patients (mean age 65) undergoing ORIF from 2015 to 2020 were chosen for this analysis.

### Variables of Interest

The primary variables of interest include patient demographics (age, sex, smoking status), comorbidities (diabetes, COPD, hypertension), and elective procedure status.

### Frailty

Frailty was measured using the RAI and mFI-5. The mFI-5 scores range from 0 to 5, with a score of 0 indicating non-frail, 1 indicating prefrail, 2 indicating frail, and scores of 3 or higher indicating severely frail. The five factors included in the mFI-5 analysis are functional dependence, diabetes, COPD, CHF, and hypertension requiring medication.^22^ The RAI evaluates a patient’s physical, cognitive, and social functions, along with relevant comorbidities such as COPD, CHF, diabetes, kidney disease, cancer, and cognitive impairments like dementia. A higher RAI score reflects a greater likelihood of adverse outcomes, with thresholds generally categorizing patients as robust, prefrail, frail, or severely frail.

Frailty was measured using the RAI and mFI-5. The mFI-5 scores range from 0 to 5, with a score of 0 indicating non-frail, 1 indicating prefrail, 2 indicating frail, and scores of 3 or higher indicating severely frail. The five factors included in the mFI-5 analysis are functional dependence, diabetes, COPD, CHF, and hypertension requiring medication. The RAI evaluates a patient’s physical, cognitive, and social functions, along with relevant comorbidities such as COPD, CHF, diabetes, kidney disease, cancer, and cognitive impairments like dementia. A higher RAI score reflects a greater likelihood of adverse outcomes, with thresholds generally categorizing patients as robust, prefrail, frail, or severely frail.

### Primary Outcome Measures

The primary outcome measures include 30-day mortality, non-home discharge (NHD), major complications (such as bleeding or wound infection), minor complications, readmission rates, and extended length of stay (eLOS > 2 days).

### Statistical Analysis

Patients were grouped based on their RAI and mFI-5 scores into categories of robust, pre-frail, frail, and severely frail (RAI only). Concordance between the RAI and mFI-5 classifications was evaluated for the same patient cohort to determine how consistently the two frailty measures aligned in stratifying patients. Demographic variables considered in the analysis included age at the time of surgery, sex, smoking history, and whether the surgery was elective. Predictive accuracy of the frailty measures was assessed using the area under the ROC curves (AUROC).27 Univariate and multivariate logistic regression analyses were performed to examine the relationship between frailty status and each outcome. Multivariate regression models controlled for confounders, including age, sex, smoking status, and elective surgery status, to ensure robust estimates of the association between frailty measures and postoperative outcomes.

Statistical analysis was performed using SPSS Statistics version 27.0.^28^ All statistical analyses used a p value < 0.05 to indicate significance. The study did not require Institutional Review Board approval due to the publicly available, de-identified nature of the NSQIP data, no consent was required from individual patients.

### Ethics Statement

This study utilized the American College of Surgeons National Surgical Quality Improvement Program (ACS-NSQIP) database, which is a de-identified, publicly available dataset. As such, this study did not involve direct interaction with human participants and did not require Institutional Review Board (IRB) approval. Participant consent was not required, and the need for informed consent was waived by the data provider due to the de-identified nature of the dataset, in accordance with institutional and national guidelines.

## Results

### Study population characteristics

The initial sample size was 4,123 patients, out of which 136 patients had incomplete data. The final sample size was 3,987 patients. The mean age was 65 (+/- 0.23) with females consisting of 62.4% [N=2574] of the study population. Tobacco use was prevalent in 18.3% [N=753] of patients. The most common comorbidities included hypertension requiring medication (37.1%) [N=1530], severe COPD (4.2%) [N=142], and diabetes requiring insulin (4.9%) or no insulin (5.1%) (Table 1.). Stratification of the study population measured by the mFI-5 included Robust: 58.7%, Prefrail: 31.1%, Frail: 9.4%, Severely frail: 0.7% (Table 2.). Stratification of the study population measured by RAI consisted of Robust: 18.3%, Prefrail: 39.5%, Frail: 35%, Severely frail: 3.9% (Table 2).

**Table 1:** Demographic and Clinical Characteristics.

| Characteristic | Number of Patients (Percentage) |
| --- | --- |
| Total patients | 4,123 (100%) |
| Gender |  |
| Female | 2,574 (62.4%) |
| Male | 1,549 (37.6%) |
| Race |  |
| White | 2,779 (67.4%) |
| Unknown/Not Reported | 1,030 (25.0%) |
| Black or African American | 168 (4.1%) |
| Asian | 127 (3.1%) |
| American Indian or Alaska Native | 14 (0.3%) |
| Native Hawaiian or Pacific Islander | 4 (0.1%) |
| Ethnicity |  |
| Non-Hispanic (N) | 2,751 (66.7%) |
| Unknown (U) | 1,082 (26.2%) |
| Hispanic (Y) | 290 (7.0%) |
| Elective Surgery |  |
| Yes | 2,545 (61.7%) |
| No | 1,570 (38.1%) |
| Unknown | 8 (0.2%) |
| Hypertension requiring medication |  |
| No | 2,593 (62.9%) |
| Yes | 1,530 (37.1%) |

**Table 2:** Frailty Indexes.

| **Frailty Index** | **Number of Patients**<br>**(Percentage)** |
| --- | --- |
| **Risk Analysis Index (RAI)** |  |
| Robust | 754 (18.9%) |
| Pre-frail | 1,628 (40.8%) |
| Frail | 1,443 (36.2%) |
| Severely frail | 162 (4.1%) |
| Missing | 136 (3.3%) |
| **5-item modified Frailty Index**<br>**(mFI-5)** |  |
| Robust | 2,422 (58.7%) |
| Pre-frail | 1,282 (31.1%) |
| Frail | 389 (9.4%) |
| Severely frail | 30 (0.7%) |

### Multivariate Analysis

Additionally, no significant findings were noted when evaluating any major complications [OR: 4.138; p=0.12], minor complications [OR: 1.270; p=0.75], readmission [OR: 0.732; p=0.63], estimated length of stay [OR: 1.072; p=0.83], or NHD [OR: 1.337; p=0.56] in the RAI frail cohorts (Table 3). The RAI severe frail cohort had a significantly increased odds for NHD [OR: 4.780; p=0.005], estimated length of stay [ OR: 2.830; p=0.008], and major complications [OR: 9.226; p=0.03] (Table 3).

**Table 3:** Multivariate Analysis Results (Risk Analysis Index)

| **Variables** | **Major complications** | **Minor complications** | **Readmission** | **Non-home discharge** | **Extended length of stay** |
| --- | --- | --- | --- | --- | --- |
| Length of total hospital stay | 1.108 (1.071 - 1.146); P<0.001 | 1.077 (1.040 - 1.115); P<0.001 | 1.042 (1.004 - 1.081); P=0.032 | 1.260 (1.213 - 1.308); P<0.001 | *** |
| Total operation time | 1.000 (0.994 - 1.005); P=0.888 | 0.999 (0.994 - 1.004); P=0.661 | 0.998 (0.994 - 1.003); P=0.467 | 0.999 (0.997 - 1.002); P=0.574 | 1.003 (1.001 - 1.005); P=0.002 |
| RAI pre-frail | 1.653 (0.404 - 6.761); P=0.484 | 0.798 (0.277 - 2.296); P=0.675 | 0.705 (0.276 - 1.796); P=0.464 | 0.542 (0.245 - 1.199); P=0.13 | 0.798 (0.487 - 1.308); P=0.371 |
| RAI frail | 4.138 (0.691 - 24.765); P=0.12 | 1.270 (0.295 - 5.466); P=0.748 | 0.732 (0.205 - 2.607); P=0.63 | 1.337 (0.509 - 3.508); P=0.556 | 1.072 (0.557 - 2.064); P=0.834 |
| RAI severe frail | 9.226 (1.251 - 68.051); P=0.029 | 2.012 (0.362 - 11.177); P=0.424 | 0.584 (0.123 - 2.766); P=0.497 | 4.780 (1.623 - 14.076); P=0.005 | 2.830 (1.312 - 6.103); P=0.008 |

| **Outcome Variable** | **Odds Ratio (95% Confidence Interval)** | **p-value** |
| --- | --- | --- |
| Non-home Discharge |  |  |
| Risk Analysis Index - Pre-frail | 0.542 (0.245 - 1.199) | 0.13 |
| Risk Analysis Index<br>- Frail | 1.337 (0.509 - 3.508) | 0.556 |
| Risk Analysis Index<br>- Severe Frail | 4.780 (1.623 - 14.076) | 0.005 |
| Age of patient<br>(patients over 89<br>coded as 90+) | 1.039 (1.022 - 1.056) | <0.001 |
| Total operation time | 0.999 (0.997 - 1.002) | 0.574 |
| Length of total<br>hospital stay | 1.260 (1.213 - 1.308) | <0.001 |
| Readmission |  |  |
| Risk Analysis Index<br>- Pre-frail | 0.705 (0.276 - 1.796) | 0.464 |
| Risk Analysis Index<br>- Frail | 0.732 (0.205 - 2.607) | 0.63 |
| Risk Analysis Index<br>- Severe Frail | 0.584 (0.123 - 2.766) | 0.497 |
| Age of patient<br>(patients over 89<br>coded as 90+) | 1.028 (1.004 - 1.052) | 0.023 |
| Total operation time | 0.998 (0.994 - 1.003) | 0.467 |
| Length of total<br>hospital stay | 1.042 (1.004 - 1.081) | 0.032 |
| Major Complications |  |  |
| Risk Analysis Index<br>- Pre-frail | 1.653 (0.404 - 6.761) | 0.484 |
| Risk Analysis Index<br>- Frail | 4.138 (0.691 - 24.765) | 0.12 |
| Risk Analysis Index<br>- Severe Frail | 9.226 (1.251 - 68.051) | 0.029 |
| Age of patient<br>(patients over 89<br>coded as 90+) | 0.999 (0.968 - 1.031) | 0.949 |
| Total operation time | 1.000 (0.994 - 1.005) | 0.888 |
| Length of total<br>hospital stay | 1.108 (1.071 - 1.146) | <0.001 |
| Minor Complications |  |  |
| Risk Analysis Index<br>- Pre-frail | 0.798 (0.277 - 2.296) | 0.675 |
| Risk Analysis Index<br>- Frail | 1.270 (0.295 - 5.466) | 0.748 |
| Risk Analysis Index<br>- Severe Frail | 2.012 (0.362 - 11.177) | 0.424 |
| Age of patient<br>(patients over 89<br>coded as 90+) | 1.006 (0.979 - 1.034) | 0.662 |
| Total operation time | 0.999 (0.994 - 1.004) | 0.661 |
| Length of total<br>hospital stay | 1.077 (1.040 - 1.115) | <0.001 |
| Extended Length of<br>Stay (>2 days) |  |  |
| Risk Analysis Index<br>- Pre-frail | 0.798 (0.487 - 1.308) | 0.371 |
| Risk Analysis Index<br>- Frail | 1.072 (0.557 - 2.064) | 0.834 |
| Risk Analysis Index<br>- Severe Frail | 2.830 (1.312 - 6.103) | 0.008 |
| Age of patient<br>(patients over 89<br>coded as 90+) | 1.032 (1.020 - 1.045) | <0.001 |
| Total operation time | 1.003 (1.001-1.005) | 0.002 |

### AUC Analysis

Analysis utilizing the AUROC was done for patients undergoing ORIF for olecranon repair in order to determine discriminatory accuracy of the mFI-5 and RAI. In regard to predicting NHD, data from RAI demonstrated superior performance [C-coefficient: 0.81; CI: 0.80 to 0.83] compared to mFI-5 [C-coefficient: 0.68; CI: 0.66 to 0.70] (p<0.001) (Table 4). Statistical differences in AUROC for reoperation was observed between mFI-5 [C-coefficient: 0.57; CI: 0.55 to 0.58] and RAI [C-coefficient: 0.63; CI: 0.62 to 0.65] (p=0.03) (Table 4). Additionally, RAI was more accurate when predicting major complications [C-coefficient: 0.72; CI: 0.70 to 0.73] compared to the mFI-5 [C-coefficient: 0.65; CI: 0.63 to 0.66] (p=0.05) (Table 4). Performances were comparable when investigating mortality between RAI [C-coefficient: 0.88; CI: 0.87 to 0.89] versus the mFI-5 [C-coefficient: 0.684; CI: 0.67 to 0.70] (p=0.08) (Table 4). No differences existed in AUROC measurements regarding minor complications for mFI-5 [C-coefficient: 0.59; CI: 0.58 to 0.61] and RAI [C-coefficient: 0.64; CI: 0.62 to 0.65] (p=0.20) (Table 4). Predictive value when assessing readmission rates when utilizing mFI-5 [C-coefficient: 0.61; CI: 0.60 to 0.63] showed no significant differences when compared to RAI [C-coefficient: 0.63; CI: 0.61 to 0.64] (p=0.63) (Table 4).

**Table 4:** Discriminatory accuracy of RAI and mFI-5 for 30-day outcomes as determined by AUROC analysis.

| Outcome | Frailty Measure | C-Statistic | 95% Confidence Interval | P-value |
| --- | --- | --- | --- | --- |
| Non-home Discharge | mFI-5 | 0.678 | 0.663 - 0.692 | <0.001 |
|  | RAI | 0.813 | 0.801 - 0.825 |  |
| Minor Complications | mFI-5 | 0.593 | 0.578 - 0.609 | 0.2001 |
|  | RAI | 0.635 | 0.620 - 0.650 |  |
| Major Complications | mFI-5 | 0.648 | 0.632 - 0.662 | 0.0499 |
|  | RAI | 0.715 | 0.700 - 0.729 |  |
| Reoperation | mFI-5 | 0.566 | 0.550 - 0.581 | 0.0336 |
|  | RAI | 0.63 | 0.615 - 0.645 |  |
| Readmission | mFI-5 | 0.613 | 0.597 - 0.628 | 0.6262 |
|  | RAI | 0.627 | 0.611 - 0.642 |  |
| Mortality | mFI-5 | 0.684 | 0.669 - 0.699 | 0.0835 |
|  | RAI | 0.878 | 0.867 - 0.888 |  |

## Discussion

Orthopedic surgeons managing olecranon fractures must consider not only the mechanical stability of the repair but also the patient’s overall physiological reserve, particularly in frail populations. The ability to predict outcomes such as non-home discharge (NHD), extended length of stay (LOS), and major complications is critical to optimizing care and resource allocation. In this study, RAI demonstrated superior performance in predicting surgical outcomes such as NHD, with an AUROC of 0.813 compared to mFI-5’s 0.678 (p<0.001). The severely frail patients, as identified by RAI, exhibited a 4.780 times increased likelihood of NHD (p=0.005), emphasizing the RAI’s ability to effectively inform discharge planning. Additionally, our findings indicate that RAI’s enhanced predictive scope can provide surgeons with a foundational basis for developing cost-effective discharge plans that mitigate unnecessary expenses and complications, particularly in the treatment of highly frail individuals within our healthcare system.^22–24^

Over the last decade, many publications in surgical literature have investigated the efficacy of various frailty tools within orthopedic settings. Indices such as the mFI-5 have been particularly prevalent in preoperative risk assessments, demonstrating consistent utility in predicting morbidity and mortality across orthopedic subspecialties.^11–14^ However, the mFI-5’s accuracy and reliability have recently been questioned due to limitations in its predictive capacity, particularly in cases involving high-frailty populations. This tool assesses frailty through five variables: functional status, diabetes history, chronic obstructive pulmonary disease (COPD) status, congestive heart failure history, and hypertension status.^15^ While the mFI-5 has demonstrated utility in preoperative risk assessment, its limited scope and equal weighting of variables reduce its sensitivity in frail populations. This tool fails to capture critical dimensions of frailty, such as cognitive impairment and functional limitations, which are included in the more comprehensive RAI. These limitations are particularly pronounced in elderly, osteoporotic patients undergoing procedures like ORIF for olecranon fractures. Indeed, Zreik et al. found that mFI-5’s predictive value for mortality was comparable to or lesser than that of the ASA classification, suggesting an area for improvement.^28^

Conversely, the Risk Analysis Index (RAI) has demonstrated considerable promise in providing a comprehensive assessment of frailty by incorporating a wider array of indicators. This 14-item index evaluates not only comorbidities and age but also impairments in activities of daily living (ADL), functional limitations, and physiological stress levels, facilitating a more robust stratification of frail patients.^16, 17^ Our analysis positions the RAI as a valuable predictor of adverse outcomes across a range of frailty levels, as evidenced by its predictive accuracy in this study, where RAI consistently outperformed mFI-5 across key outcomes, including non-home discharge (NHD), reoperation, and major complications. Despite its recent validation within surgical settings, relatively few studies have applied the RAI specifically to moderate-risk surgeries like ORIF for olecranon fractures. Our study is therefore unique in this context, examining the implications of frailty in olecranon fracture cases through both the RAI and mFI-5 lenses.

Demographic and clinical factors, including advanced age (>80 years) and female sex, were found to significantly influence NHD rates among patients undergoing ORIF for olecranon fractures. The RAI’s comprehensive framework, which includes frailty dimensions not covered by mFI-5, provides a more granular view of physiological vulnerabilities, particularly in elderly female patients who disproportionately experience osteoporotic fractures.^25^ Our analysis found that the RAI consistently outperformed mFI-5 in predicting adverse outcomes across frail and severely frail cohorts, with RAI demonstrating significantly higher AUROC values (p<0.001). These results suggest that RAI’s broader health assessment parameters—incorporating cognitive impairment, functional limitations, and expanded comorbidity profiling—allow for more precise stratification of risk. This predictive advantage directly informs discharge planning by identifying patients who may require intensive post-acute care, especially among severely frail elderly females.^26, 27^ The enhanced predictive capacity of the RAI compared to the mFI-5 provides orthopedic surgeons with actionable insights for planning perioperative and postoperative care. For instance, patients identified as severely frail by the RAI may benefit from targeted interventions such as prehabilitation, enhanced monitoring, or early engagement of rehabilitation services. Additionally, the RAI’s ability to stratify risk in elderly, osteoporotic females—a population disproportionately affected by olecranon fractures—underscores its clinical utility in tailoring care strategies to individual patient profiles.

Frail patients present unique challenges in the context of olecranon fracture repair, including poor bone quality, slower rehabilitation progress, and higher risks of hardware failure or nonunion. These factors are compounded by the need for multidisciplinary perioperative management, especially in elderly individuals with osteoporotic fractures or concurrent injuries. Our findings suggest that integrating the RAI comprehensive frailty assessment into preoperative workflows can aid in anticipating these challenges and mitigating associated risks. Within our patient cohort, higher rates of NHD were observed as RAI scores increased, underscoring the tool’s relevance in guiding postoperative planning in cases where frail patients are more likely to experience severe fractures. These findings align with our observation that olecranon fracture patients with elevated RAI scores often have concurrent injuries, such as hip fractures, which further complicate the recovery process.^19, 20^ Previous literature on moderate-intensity orthopedic surgeries has shown that elevated RAI scores correspond with increased rates of adverse events, a trend our data reinforce, particularly within the severely frail cohort.^21^

RAI’s inclusion in preoperative assessments could optimize the allocation of post-acute care resources, reducing the high costs associated with prolonged length of stay (LOS) and readmissions often seen in frail elderly populations. Within this study, RAI’s predictive accuracy was most evident in the severely frail cohort, where a marked increase in major complications (OR: 9.226; p=0.03) and extended LOS (OR: 2.830; p=0.008) was observed. This pattern reinforces the RAI’s sensitivity to a broader set of frailty indicators, beyond those included in mFI-5, and highlights the index’s relevance for capturing complex patient profiles. By accurately identifying high-risk cases in advance, the RAI can support a refined approach to perioperative risk assessment, facilitating better-informed patient transitions to post-acute care settings, and ultimately improving both resource efficiency and patient outcomes.^28^ Given the financial pressures associated with prolonged hospital stays and readmissions, incorporating the RAI into standard preoperative protocols has the potential to reduce unnecessary expenditures. This is particularly relevant in value-based care models, where accurate risk stratification enables orthopaedic departments to target resource allocation, such as referral to skilled nursing facilities or intensive rehabilitation services, for patients at highest risk.^23–26^

The observational nature of this study may impede its ability to determine causality between variables. Additionally, data from NSQIP did not record variables such as time of injury to operation, severity of fracture, or fracture type. Analysis from the study did not explore the presence of other comorbid fractures that may have been present that could possibly require additional operations. Also, the RAI may lead to overestimation in very frail patients, or need to be recalibrated across diverse patient populations. Despite these limitations, the robust nature of the statistical findings and data from the ROC curve remain reliable. Additionally, the large sample size of 3,987 patients provides adequate power for our analysis. Future studies should explore additional variables, such as injury-to-operation timing and fracture severity, to enhance the predictive power of frailty tools. Furthermore, recalibrating the RAI for diverse patient populations will ensure its applicability across different clinical contexts.

## Conclusion

The RAI demonstrated superior discriminatory accuracy when investigating NHD and major complications when compared to the mFI-5. Additionally, the adverse effects of ORIF in olecranon fractures were more prevalent in severely frail cohorts as opposed to pre frail and frail cohorts, suggesting the implementation of frailty scores would better serve surgeons when determining the appropriate intervention. Indeed, this study underscores the importance of incorporating the RAI frailty assessment tool into the clinical management of olecranon fractures in frail populations. By improving the accuracy of outcome predictions, the RAI not only aids surgeons in preoperative planning but also enhances postoperative care strategies, ultimately leading to better patient outcomes and more efficient use of healthcare resources.

## Data Availability

The data underlying the results presented in this study are available from the American College of Surgeons National Surgical Quality Improvement Program (ACS-NSQIP). The dataset is available to researchers through a formal application process at https://www.facs.org/quality-programs/data-and-registries/acs-nsqip/participant-use-data-file/. The authors do not have permission to share the dataset directly.

## Acknowledgments

None

---

**Figure 1A.**
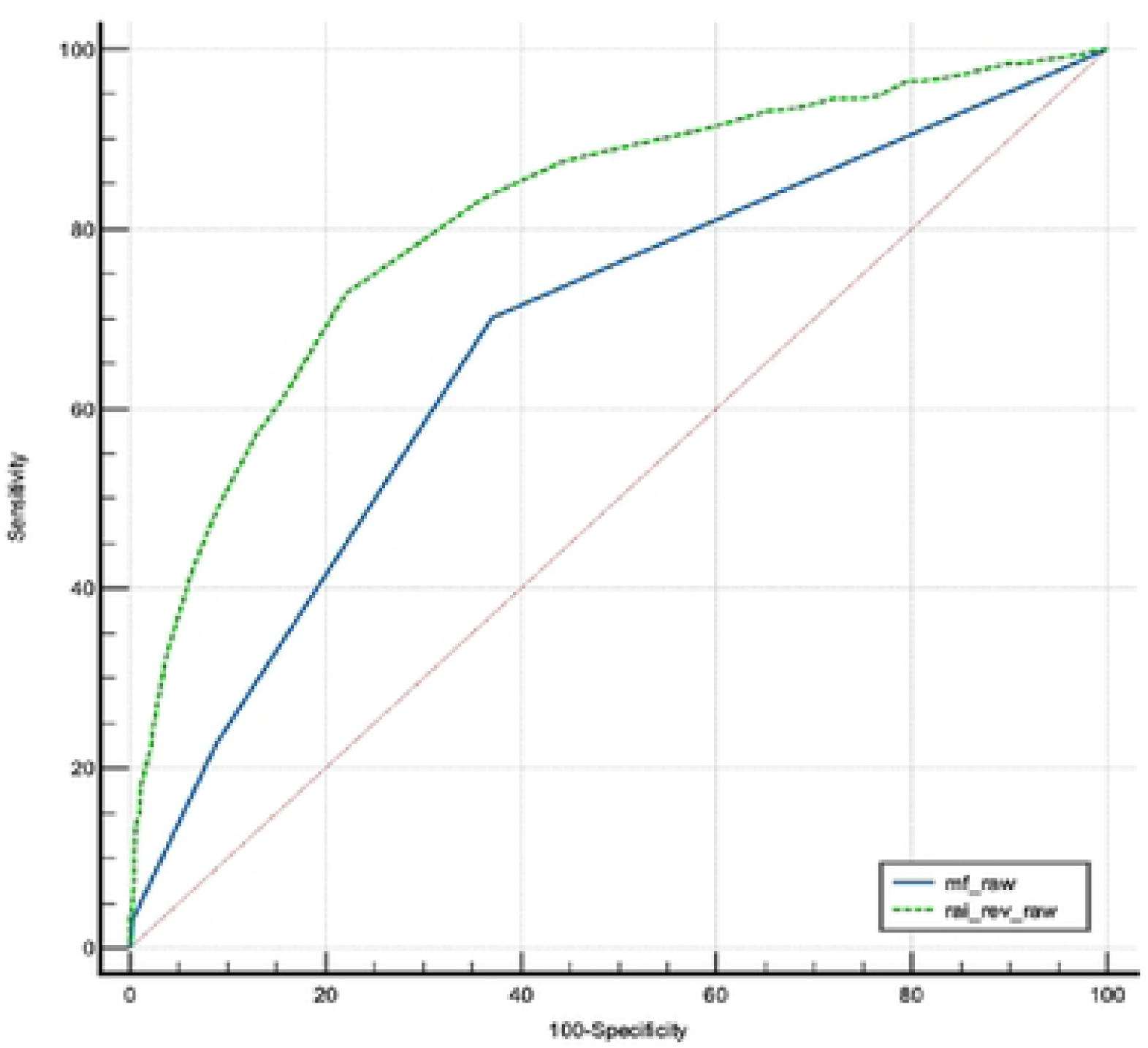
RAI vs mFl-5 for Non-home discharge.

**Figure 1B.**
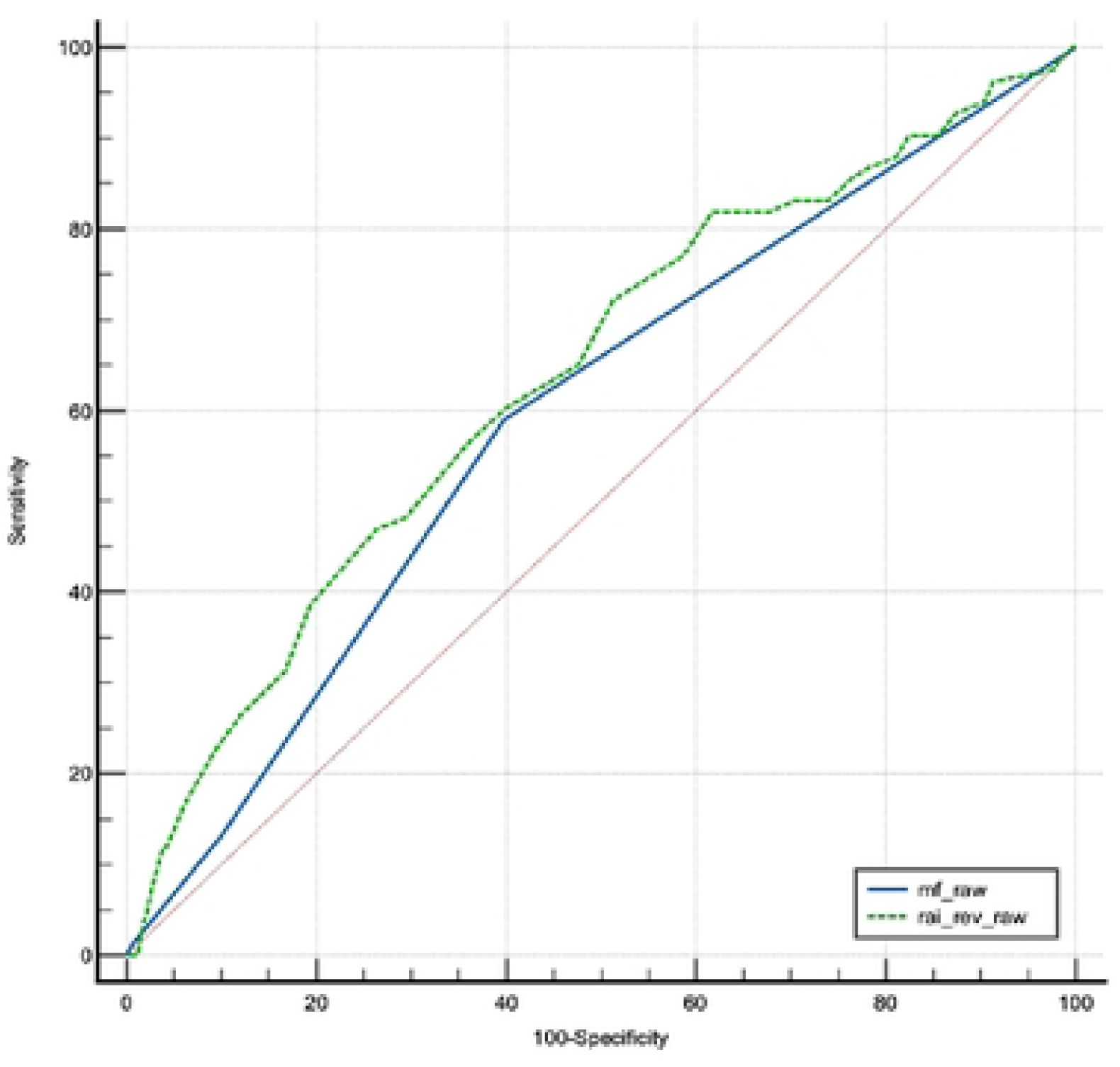
RAI vs mFI-5 for Minor complications.

**Figure 1C.**
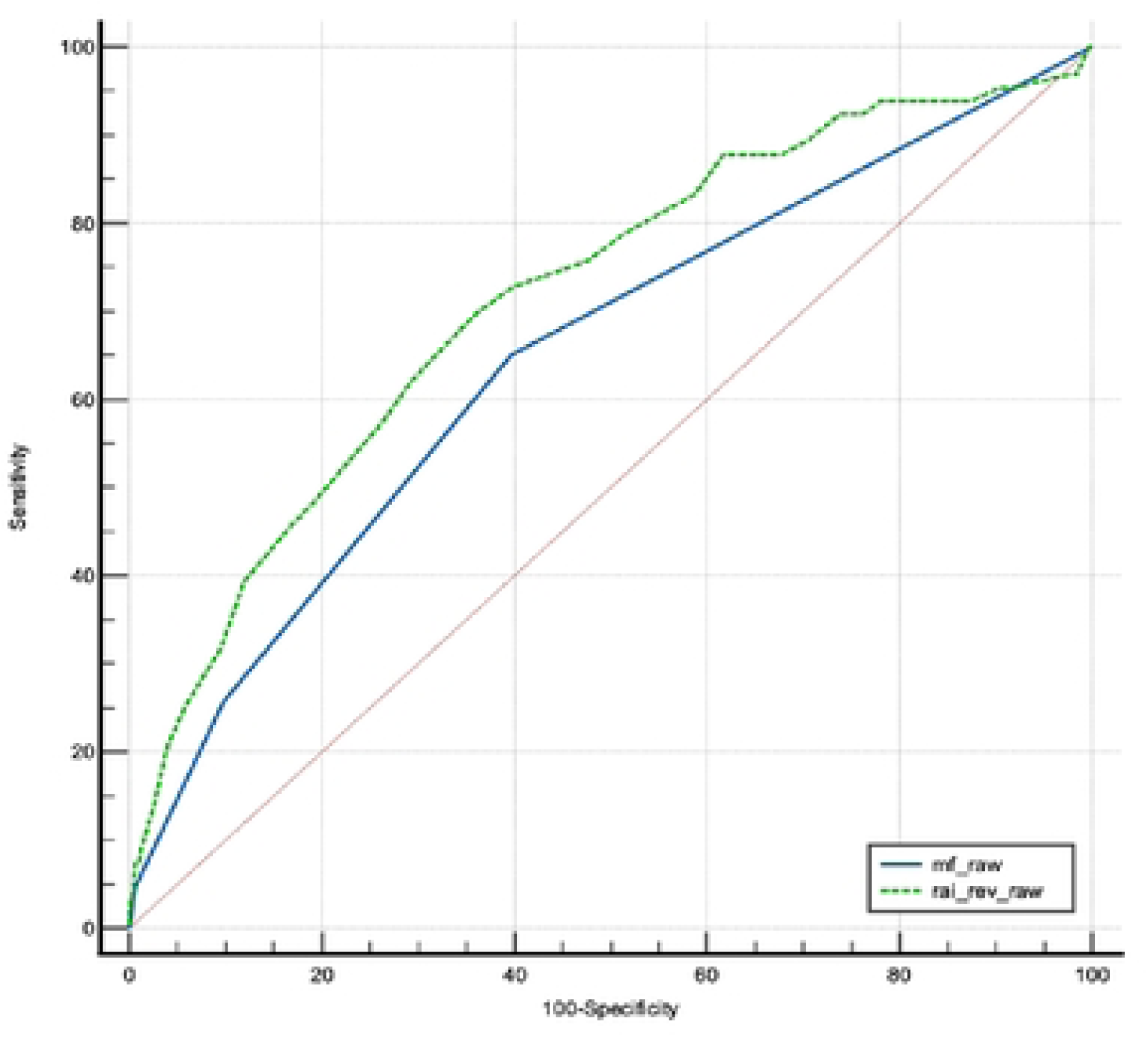
RAI vs n1FI-S for Major complications.

**Figure 1D.**
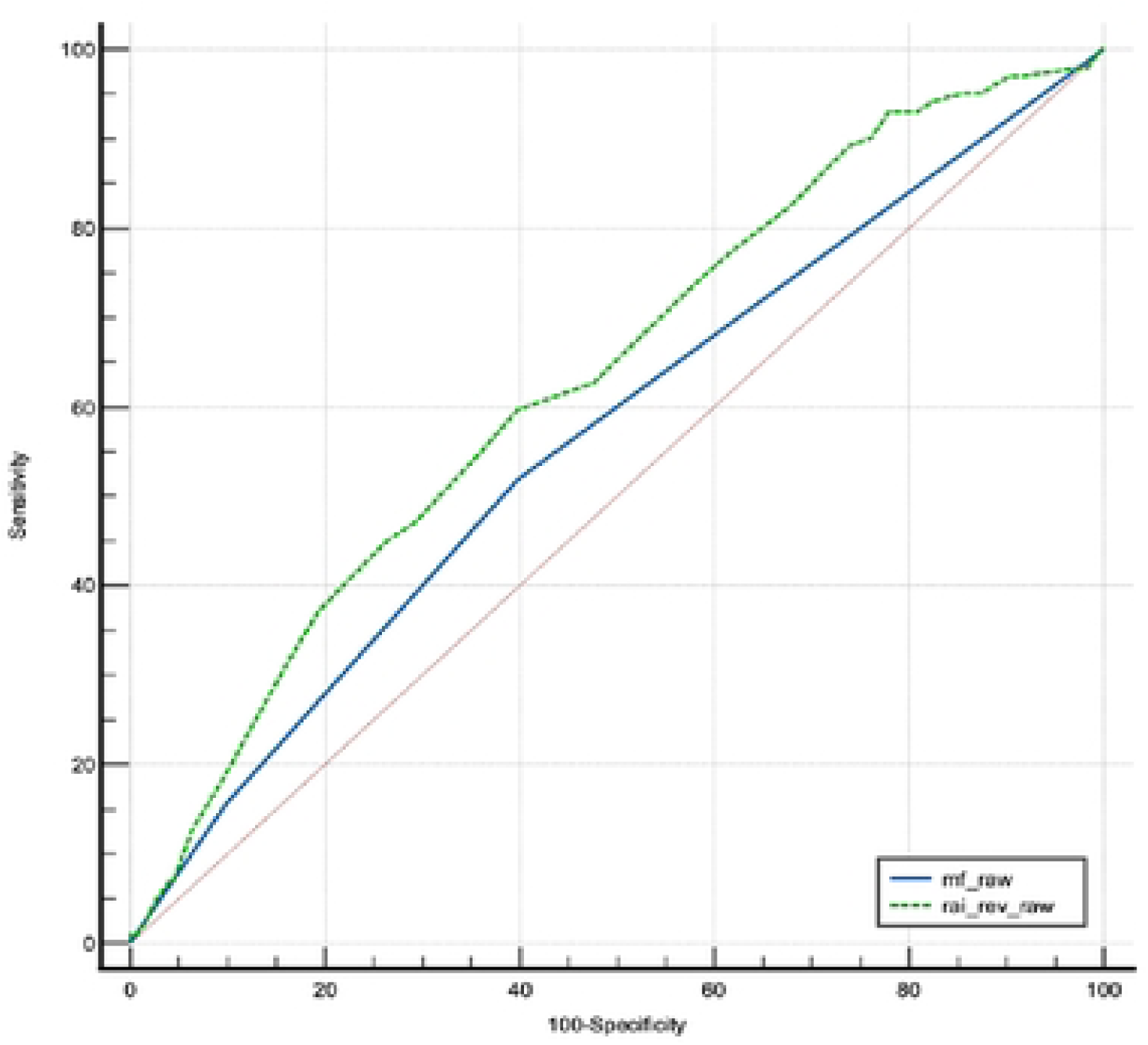
RAI vs n1FI-S for Reoperation.

**Figure 1E.**
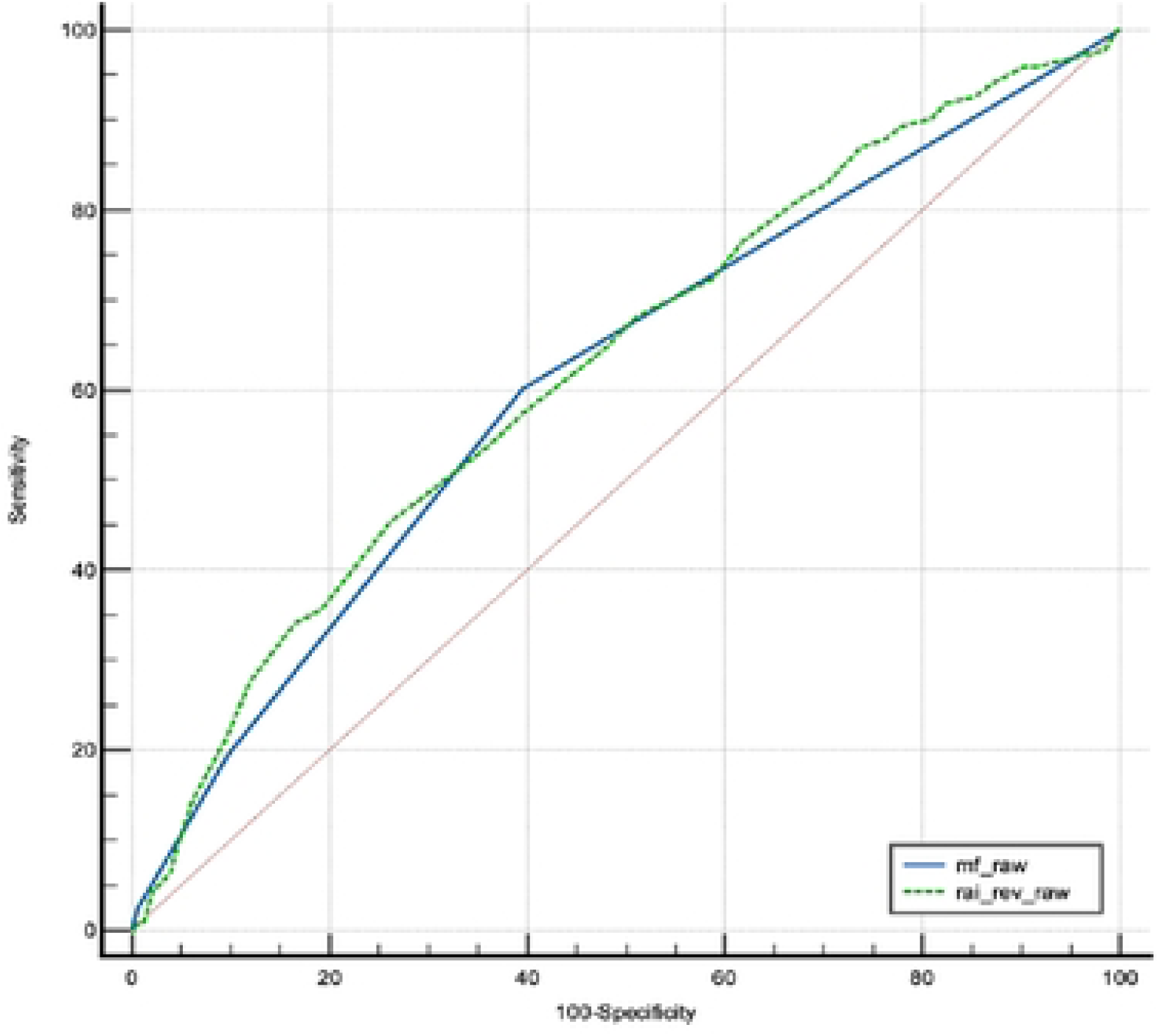
RAI vs mFI-5 for Readmission.

**Figure 1F.**
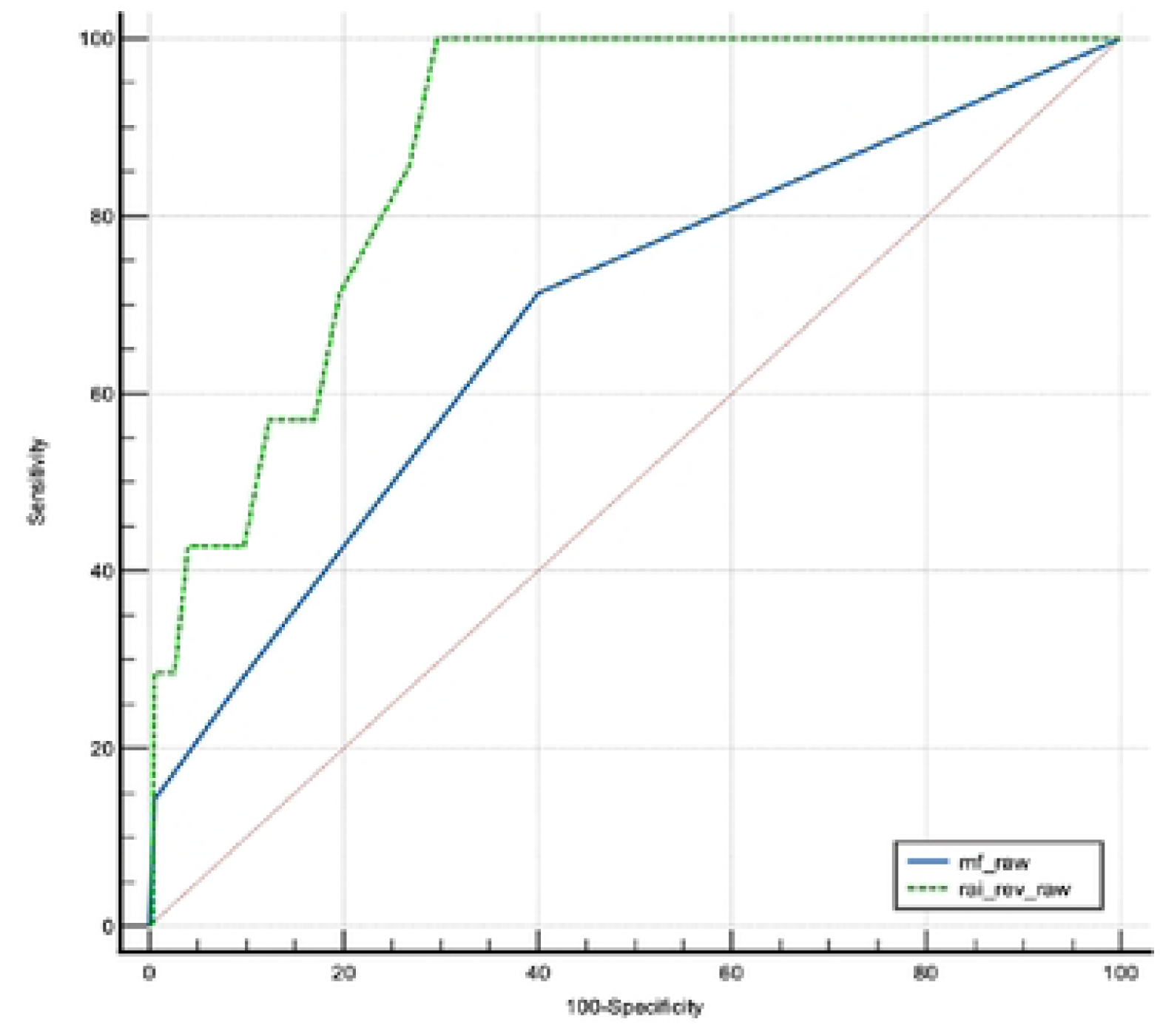
RAI vs mFI-5 for Mortality.

